# Is sleep apnea-hypopnea index relevant for impaired brain perfusion and desaturation in patients with severe obstructive sleep apnea syndromes?

**DOI:** 10.1101/2020.10.14.20212639

**Authors:** Zhongxing Zhang, Ming Qi, Gordana Hügli, Ramin Khatami

**Affiliations:** Center for Sleep Medicine and Sleep Research, Clinic Barmelweid AG, Barmelweid, Switzerland; Department of Neurology, Inselspital, Bern University Hospital and University of Bern, Bern, Switzerland

## Abstract

**Background:** Obstructive sleep apnea syndrome (OSAS) is a common sleep disorder with the prevalence of 9-38% in the general population. Severe OSAS defined as apnea-hypopnea index (AHI) ≥ 30/h is a major risk factor for developing cerebro-cardiovascular diseases like stroke. Treatments such as continuous positive airway pressure (CPAP) therapy are recommended for those patients because untreated OSA can cause tremendous medical expenses and economic burden. Understanding the mechanisms of how repetitive sleep apneas/hypopneas induce changes in cerebral hemodynamics in severe OSAS is crucial for the understanding of disease severity, disease progression in brain damage and the treatment efficacy. However, these mechanisms and the association between AHI and the induced changes in cerebral hemodynamics in severe OSAS are essentially unknown. Using a stepwise incremental CPAP titration protocol we aim to identify the most relevant physiological factors including AHI and their quantitative contributions to cerebral hemodynamic changes before and during CPAP treatment in severe OSAS patients.

**Methods and findings:** 29 newly diagnosed severe OSAS patients underwent incremental CPAP titration during polysomnography recordings: 1-h sleep without CPAP served as self-controlled baseline followed by stepwise increments of 1-cmH2O pressure per-hour starting from 5-8 cmH2O individually. Frequency-domain near-infrared spectroscopy measured the changes in blood volume (BV) and oxygen saturation (StO2) in the left forehead. The coefficients of variation of BV (CV-BV) and the decreases of StO2 (de-StO2) during more than 2000 respiratory events were predicted by various predictors including AHI using linear mixed-effect models, respectively. The best predictors were automatically selected by stepwise regression. Surprisingly, AHI was excluded from the final selected models. Longer events and apneas rather than hypopneas induce larger changes in CV-BV and stronger cerebral desaturation. Respiratory events occurring during higher baseline StO2 before their onsets, during rapid-eye-movement sleep and those associated with higher heart rate (HR) during the events trigger smaller changes in CV-BV and de-StO2. CPAP pressures attenuate the changes in CV-BV and de-StO2.

**Conclusions:** In severe OSAS the length and the type of respiratory event rather than widely used AHI may be better parameters to indicate the severity of cerebral vascular damage. Thus both, length and type of respiratory events should be considered as predictors of cerebro-cardiovascular events in future epidemiological studies and in clinical practice. OSAS patients with profound long apnea events need to pay special attention and quickly treated with CPAP therapy as it can restore the induced cerebral hemodynamic changes.

## Introduction

Obstructive sleep apnea syndrome (OSAS) is a common sleep disorder with the prevalence of 9-38% in the general population [1, 2]. Currently, the severity of OSAS is classified by the apnea-hypopnea index (AHI) which is the number of apnea and hypopnea events per hour during sleep [3]: 5≤AHI<15 is mild, 15≤AHI<30 is moderate and AHI≥30 is severe. Epidemiological studies have repeatedly confirmed that the severity of OSAS plays a significant role in the development of cerebrovascular diseases including stroke [4-7], although the cut-off values of AHI are different in these studies. For example, Redline et al. reported that OSAS patients with AHI ≥20 in men and ≥ 25 in women have significantly increased risk of stroke compared with those without OSAS (i.e., AHI<5) [4]. The similar cut-off of AHI≥20 is associated with increased risk of stoke is also founded by Arzt et al. [5] whereas cut-off values given by Yaggi et al. [6] and Bassetti et al. [7] are AHI≥35 and ≥30, respectively. Nevertheless, patients with severe OSAS are at high risk of developing stroke.

The current classification of SAS severity carries two problems for patients with cerebrovascular disease. First, an AHI ≥30 contains a broad range of numbers as some patients’ AHI can be higher than 100. Whether in severe OSAS AHI can further correlate to the risk of cerebrovascular diseases is still unknown. Second, in severe OSAS previous studies found that the recurrent respiratory events can induce repetitive changes in cerebral perfusion and oxygenation [8-14], which lead to reduced cerebrovascular reactivity and increased arterial stiffness [8]. These studies suggested that OSAS patients chronically exposed to high frequent (i.e., high AHI) repetitive hypoxemia and cerebral perfusion changes are at permanent risk to cerebral ischemia, because the endothelium and the cerebral autoregulation (CA) may be eventually damaged. However it remains unknown if the underlying mechanisms of cerebrovascular damage are indeed reflected by the AHI. Intact CA during both wake and sleep is essential to maintain a constant perfusion of brain tissue and a prerequisite to prevent acute and chronic brain damage [14-16]. Indeed, a recent study demonstrated recurrence of cerebral deoxygenation along with recurrence of previously treated nocturnal breathing disturbances when therapeutic CPAP was withdrawn [12]. Thus, a better understanding of how recurrent sleep apneas/hypopneas affect CA may provide valuable insights into the acute and chronic consequences including cerebral damage of OSAS [8-10, 17]. However, although previous studies [8-14] could demonstrate that repetitive changes in cerebral deoxygenation and perfusion are directly caused by the respiratory events, the exact mechanisms are mainly determined by AHI or by other mechanisms, is still essentially unknown.

Change in cerebral hemodynamics is a net effect resulting from a combination of multiple pathophysiological factors that challenge CA in OSAS, e.g., cerebral deoxygenation, cardiac output and heart rate (HR), intrathoracic pressure (ITP) swings, hypercapnia, hypoxia, sleep stages and arousals. For example, the ITP swings during OSAS events are associated with decreased cardiac output and HR [18-20], leading to a reduced global blood/oxygen supply to the brain. On the other hand, acute hypoxia and hypercapnia cause an increase in cerebral blood flow via dilations of cerebral arteries and arterioles [21], counteracting the increase in blood pressure and blood supply. In addition, sleep related factors such as different sleep stages [22-24], and arousals can also cause changes in cerebral hemodynamics [25, 26], adding to the complexity of cerebral regulation during sleep.

In this study we aim to investigate how the aforementioned different pathophysiological mechanisms quantitatively affect changes in cerebral perfusion and oxygenation in severe OSAS. We used near-infrared spectroscopy (NIRS), a well-validated optical technique in assessing the cerebral hemodynamic changes in apneas/hypopneas [10-13, 16, 27, 28], to measure the local hemodynamic changes induced by apneas/hypopneas in microvascular bed which is more challenging for CA [29, 30]. NIRS is sensitive to hemodynamic changes in microvascular bed (i.e., arterioles, venules and capillaries) [31, 32]. It can simultaneously measure changes in both blood volume (BV) and tissue oxygen saturation (StO2), thus providing better insights into the cerebral hemodynamic consequences of OSAS. We hypothesize that AHI may be not the only factor determining the degree of cerebral hemodynamic changes induced by apneas and hypopneas considering the aforementioned complexity of cerebral hemodynamic regulations; but it may be the key factor considering that it currently defines the severity of OSAS.

## Methods

Totally 29 newly diagnosed OSAS patients (age [mean ± standard deviation, std.]: 54.7±13.9 years, interquartile range [IQR] 42-65 years; male: 26; body-mass-index [BMI]: 35.8±7.2, IQR 31.9-41.6; apnea-hypopnea-index [AHI]: 53.1± 24.6/h, IQR 32.7-70 /h) participated in this study. Patients with unstable coronary or cerebral artery disease, severe arterial hypertension or hypotension, respiratory diseases or a history of a sleep-related accident were excluded. This study was approved by the Kantonale Ethikkommission Aargau, Switzerland, and was in compliance with the declaration of Helsinki.

In the first night all the patients did video-polysomnography (PSG) measurement (Embla RemLogic, Natus Medical Incorporated, Tonawanda, NY, USA) for diagnosis. On next day the patients diagnosed as OSAS and clinically recommended for CPAP therapy gave their written informed consent. In the following night they did the stepwise CPAP (AirSense™10, ResMed) titration together with video-PSG and NIRS recordings: 1-h sleep without CPAP served as the self-controlled baseline followed by stepwise increment of 1-cmH2O pressure per-hour starting from 5-8 cmH2O depending on the individuals.

video-PSG measured electroencephalography at electrode locations of C3, C4, O1, O2, F3, and F4 according to 10–20 system, electrooculogram, electromyogram, electrocardiogram, breathing functions, HR, peripheral oxygen saturation and the movement during sleep. Two experienced sleep technologists independently scored the sleep stages, respiratory and limb movement events, and motion artifacts in 30-s epochs according to the 2017 American Academy of Sleep Medicine manual [3]. The discrepancy between these two technologists was solved by their discussion or the recommendation of the third experienced neurophysiologist. The hourly AHI, arousal-index (AI), leg-movement-index (LMI) under specific CPAP pressure per-hour were also calculated (i.e., the number of events divided by the sleep time under each CPAP pressure per-hour in the titration protocol).

Frequency-domain multi-distance (FDMD)-NIRS (Imagent, ISS, Champaign IL, USA) measurements were conducted over the middle of left forehead. The cerebral oxygenated (HbO2) and deoxygenated (HHb) hemoglobin were calculated from the fitted slopes of the modulated light amplitude and phase shifts over four various light sources and detector distances [33, 34]. The sum of HbO_2_ and HHb was the cerebral BV. The absolute value of StO2 was also calculated in FDMD-NIRS [34]. The sample rate of FDMD-NIRS recording was 5.2 Hz. 2-s preceding and 5-s following the sleep apnea/hypopnea event was selected as baseline and recovery phase, respectively. More details of the FDMD-NIRS measurement can be found in the supplemental file.

The coefficient of variation (CV) of BV (CV-BV) during apnea/hypopnea and recovery phase was calculated. CV is a standardized measure (i.e., standardization) of the variability of the data in relation to their mean, i.e., CV=100×std./mean. It was used in previous NIRS studies quantifying the cerebral hemodynamic changes between subjects as it can eliminate the bias due to individual differences [32, 35]. Smaller changes in CV-BV indicated smaller changes in the cerebral perfusion. The mean StO2 at baseline before event onset and the subsequent decrease (de-StO2, which was the maximal value minus the minimal value after event onset) were also calculated.

Linear mixed-effects model (LMM) with a random intercept by patients was used to quantify which parameters determining the CV-BV and the de-StO2 caused by the apneas/hypopneas, respectively. The explanatory variables were age, sex, BMI, sleep stages, sleep positions, types and durations of respiratory events, mean HR during the events, the baseline StO2 before the onset of the event, CPAP pressures and the per-hour AHI/AI /LMI under each pressure. In each patient all the events under a specific CPAP pressure were excluded from LMM if the corresponding sleep duration under that pressure was shorter than 20 minutes, to exclude the unreliable calculation of the per-hour AHI/AI/LMI. In total the data of 2167 events from 27 patients were selected. One male was excluded because of the falling off of his position sensor. One female was excluded because of short sleep time. Box plot was used to graphically depict the CV-BV and the de-StO2. Outliers of the box plots were further excluded from LMM to avoid violating the model assumptions of normality and homoscedasticity. Stepwise regression using both forward selection and backward elimination was done to select the best predictors. We reported both the conditional *R*^*2*^ [36] and Ω^*2*^ [37] to assess the goodness of fit of our final selected models.

Data were expressed as the mean ± standard error (SE) unless otherwise indicated. All statistical analyses and modeling were performed using R (version 3.2.4). The LMM models were done using the R package *lme4* and *lmerTest*.

## Results

### Changing trends of cerebral hemodynamics

In total FDMD-NIRS successfully measured 884 events (205 obstructive apneas and 679 hypopneas) at 1-hour baseline recordings without CPAP and 1873 events (239 obstructive apneas and 1634 hypopneas) during stepwise CPAP titrations. The distributions of the durations of the events at baseline and during titration were shown in Fig.1. The most frequent events were 12-16 seconds at baseline and 13-20 seconds during CPAP titration.

**Fig.1.**
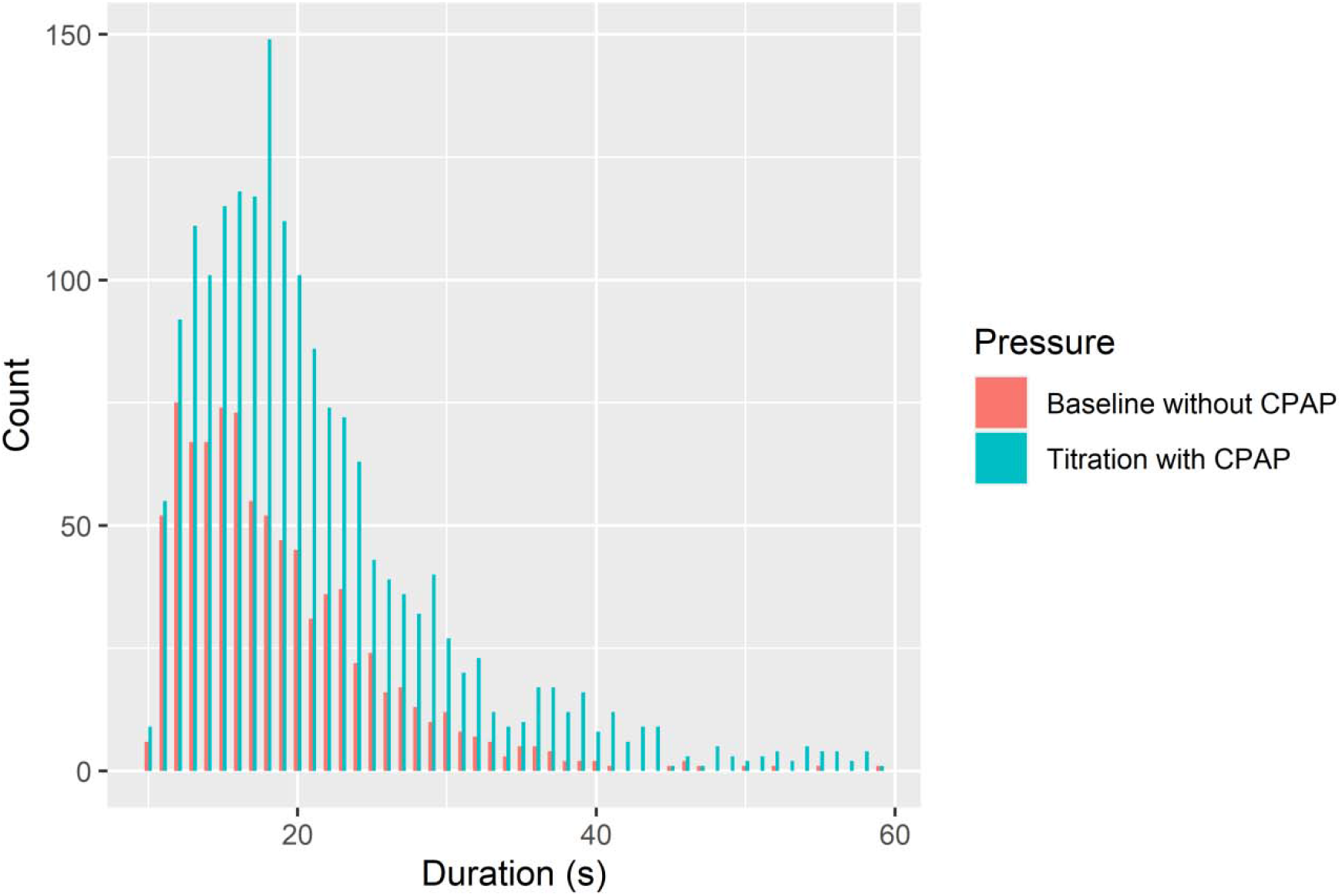
The histogram of the durations of the respiratory events under the measurements without CPAP (n=884) and with the CPAP (n=1873). 55 events longer than 60 seconds during titration are not shown in the figure.

Fig.2 showed a sample of NIRS changes. Cerebral HbO2 and StO2 decreased and HHb increased after the onsets of the events and their changes were reversed after the end of the events, which was a typical hemodynamic pattern triggered by obstructive apneas and hypopneas reported in previous studies [10, 11, 13, 28]. There was a phase shift between BV and StO2. BV started to decrease before the event onsets and then increased after reaching its nadir within the events, while StO2 started to decrease after the event onsets and it reached its nadir later than BV. The increased BV within the events could be due to cerebral vasodilation triggered by acute hypoxia and hypercapnia. The decrease in BV that occurred after the end of the events could indicate cerebral vasoconstriction in the recovery phase.

**Fig.2.**
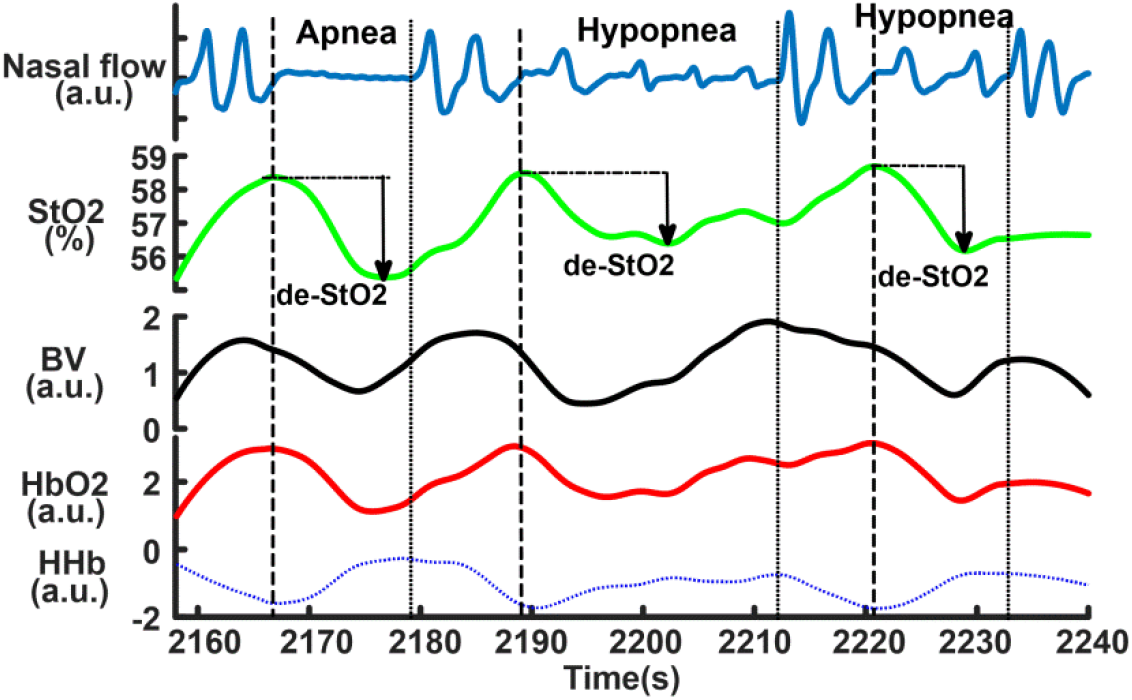
The samples of NIRS changes in obstructive sleep apneas and hypopneas. The dash lines indicate the start and the end of the events. The cerebral desaturations (de-StO2) are marked in the three events. BV is blood volume. a.u. is arbitrary unit.

## Results of LMMs analyses

The CV-BV and de-StO2 were significantly correlated as shown in Fig.3 (A). The distribution of the CV-BV was shown in Fig.3 (B) and its mean value was 1.4± 0.02% (IQR: 0.7%-1.6%). Its maximum was 3% as shown in the box plot in Fig. 3 (B), so the data larger than 3% were outliers (n=156). Similarly, the distribution of the de-StO2 (mean value: 4.02±0.06%, IQR: 2.15%-4.79%) was illustrated in Fig.3 (C) and box plot suggested that the data larger than 8.7% were outliers (n=149). Thus 2011 (247 obstructive apneas and 1764 hypopneas) and 2018 (248 obstructive apneas and 1770 hypopneas) events were finally used to build the LMMs for the CV-BV and the de-StO2, respectively.

**Fig.3.**
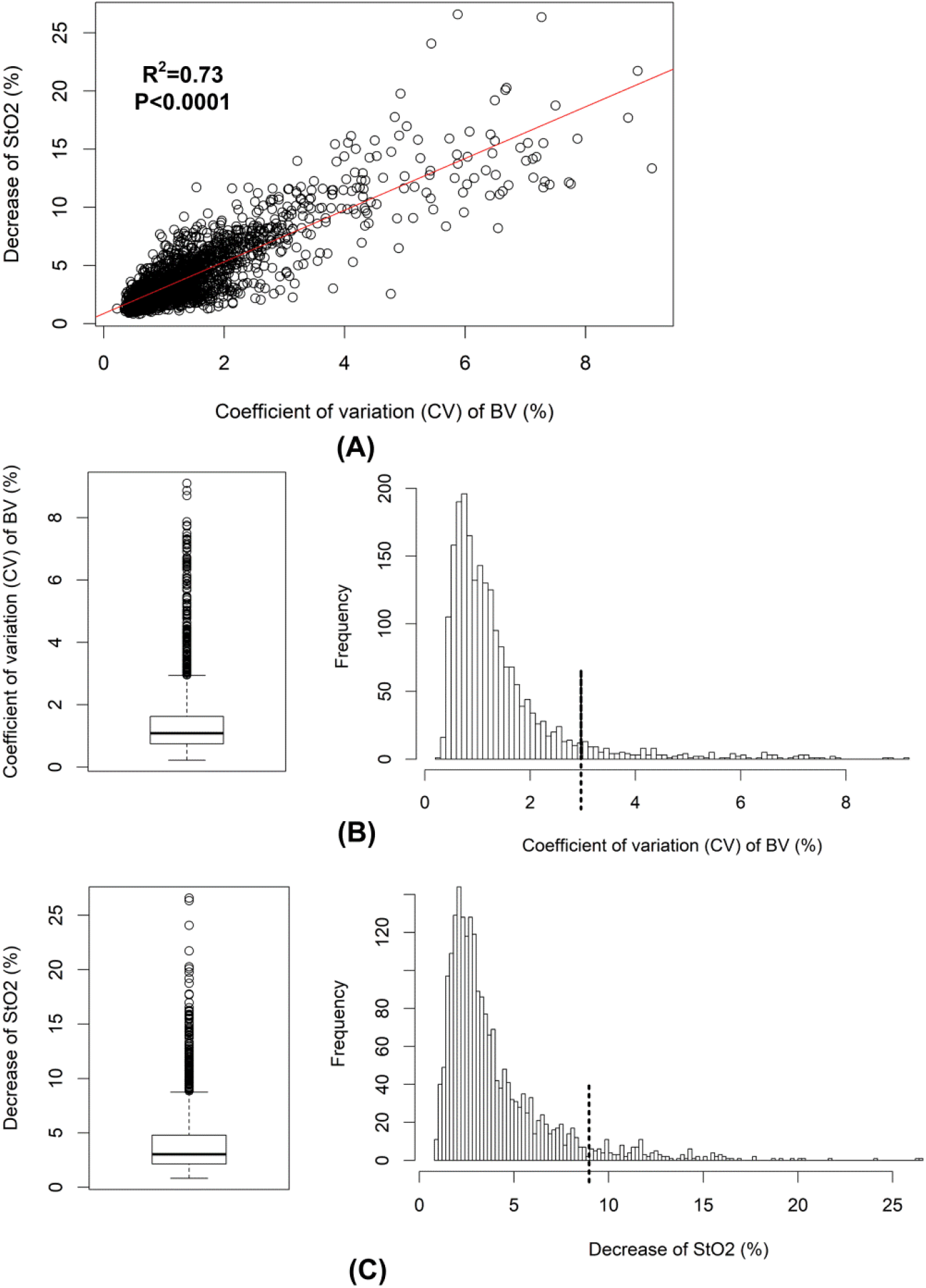
The distributions of the coefficient of variation (CV) of cerebral blood volume (BV) and the decrease of cerebral StO2 (de-StO2). Changes in CV of BV and de-StO2 are correlated as shown in (A). The box plot suggests the outliers of CV of BV are larger than 3% and the cut-off 3% is marked with the dash line in its distribution (B). Similarly, the cut-off for outliers is 8.7% as shown in the box plot of de-StO2 and it is marked with the dash line in the distribution (C).

The outcomes of the final LMMs selected by stepwise regressions were shown in Table 1 and Table 2. The conditional *R*^*2*^ and Ω^*2*^ of the model for CV-BV were 0.77 and 0.65, respectively. These two values of the model for de-StO2 were 0.81 and 0.72, respectively. In both models age, sex, BMI, AHI at diagnosis, and per-hour AHI/LMI were eliminated by the stepwise selections. The results were summarized as:

**Table 1.** The outcomes of the linear mixed-effects model of the CV-BV changes.

|  | Estimate | Std. Error | t-value | P-value |
| --- | --- | --- | --- | --- |
|  | (10<sup>-3</sup>) | (10<sup>-3</sup>) |  |  |
| Duration of events | 2.66 | 0.57 | 4.67 | <0.0001 |
| Apnea-Hypopnea | 195.3 | 28.97 | 6.74 | <0.0001 |
| Mean HR within events | -5.16 | 1.22 | -4.24 | <0.0001 |
| Hourly arousal index | 2.14 | 0.45 | 4.8 | <0.0001 |
| Baseline StO2 | -10.77 | 3.22 | -3.34 | 0.00085 |
| REM sleep-NREM sleep | -64.75 | 29.95 | -2.16 | 0.031 |
| Sleep positions |  |  |  |  |
| Right side | 172.5 | 34.65 | 4.98 | <0.0001 |
| Supine | 130.0 | 31.16 | 4.17 | <0.0001 |
| CPAP pressures | -5.93 | 2.54 | -2.33 | 0.0198 |

HR: heart rate. REM: rapid eye movement. NREM: non-REM. CPAP: continuous positive airway pressure. The change in apnea is the reference for the change in hypopnea in this model. Sleep on left side is the reference for sleep on right side and on supine position. NREM sleep is the reference for REM sleep.
Hourly arousal index is the arousal index calculated in each hour.

**Table 2.**
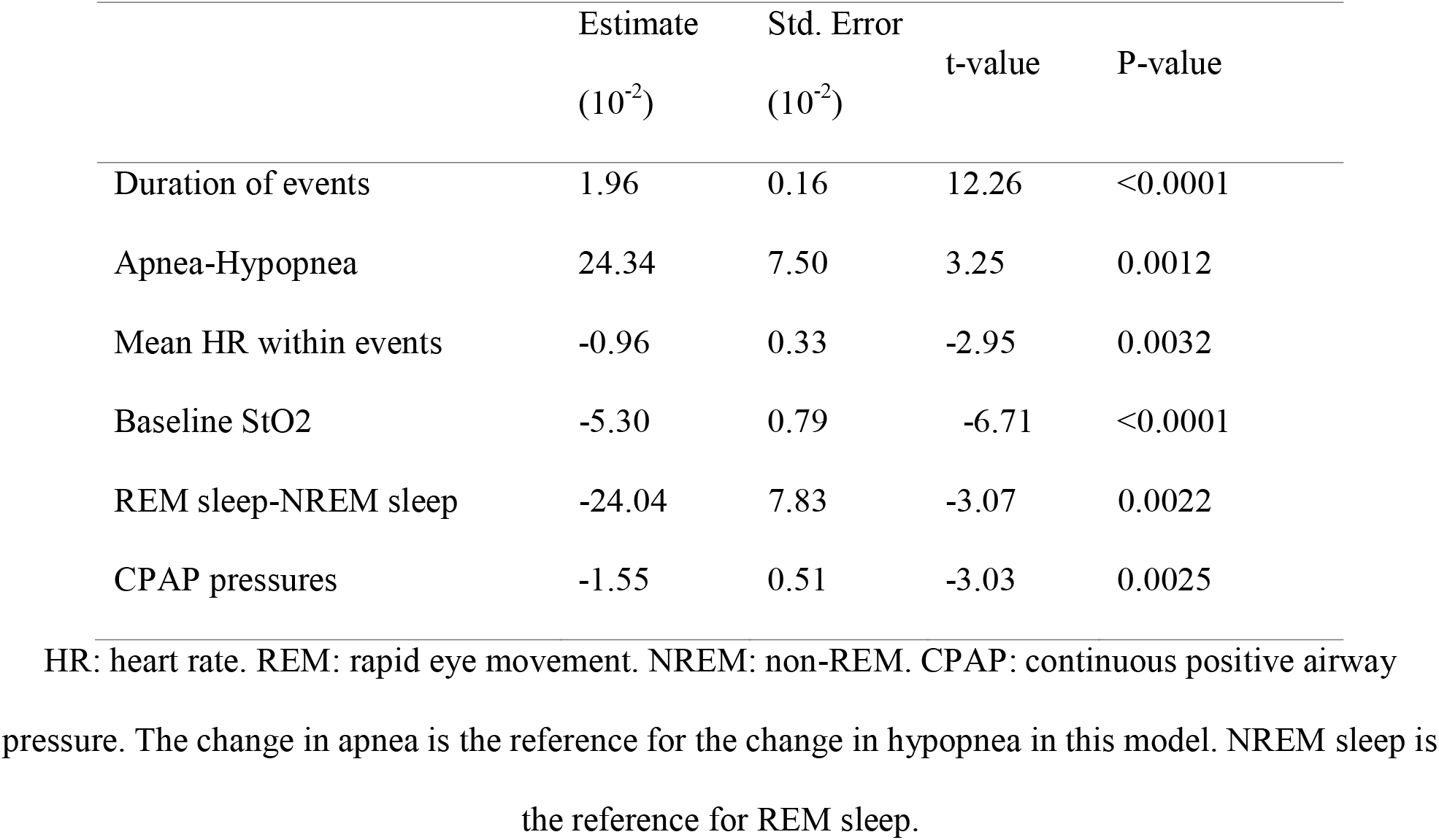
The outcomes of the linear mixed-effects model of the decrease of StO2.

1. The duration and the type of the events were the most significant covariates determining the changes in cerebral hemodynamics. The changes in cerebral perfusion (i.e., CV-BV) and cerebral desaturation (i.e., de-StO2) were larger in the events of longer durations after the other covariates were controlled. Obstructive apneas induced larger CV-BV and cerebral desaturation compared to hypopneas.
2. Events associated with larger mean HR and higher baseline StO2 induced smaller changes in cerebral perfusion and desaturation.
3. Events in rapid eye movement (REM) sleep caused smaller changes in CV-BV and de-StO2 compared to the ones in non-REM (NREM) sleep.
4. Arousal system influenced the induced changes in cerebral perfusion but not in cerebral desaturation.
5. Sleep position did not affect cerebral desaturation but only the perfusion. The events caused larger changes in CV-BV when the patients slept on their right side or on their back compared to sleep on the left side. There was no difference in CV-BV between sleeping on the right side and on the supine position (stepwise regression gave t-value=1.60, P-value=0.11).
6. Finally, increased CPAP pressure can stepwise attenuate the event-related changes in cerebral hemodynamics.

Since it was unexpectedly to find that the event’s length rather than AHI was the most significant predictor of the changes in cerebral hemodynamics, we illustrated the changes in CV-BV and de-StO2 versus the event’s duration in each patient in Fig.4. The individual linear fitting lines in Fig.4 suggested that indeed in the majority of the patients the changes in CV-BV and de-StO2 increased with the increases in the durations.

**Fig.4.**
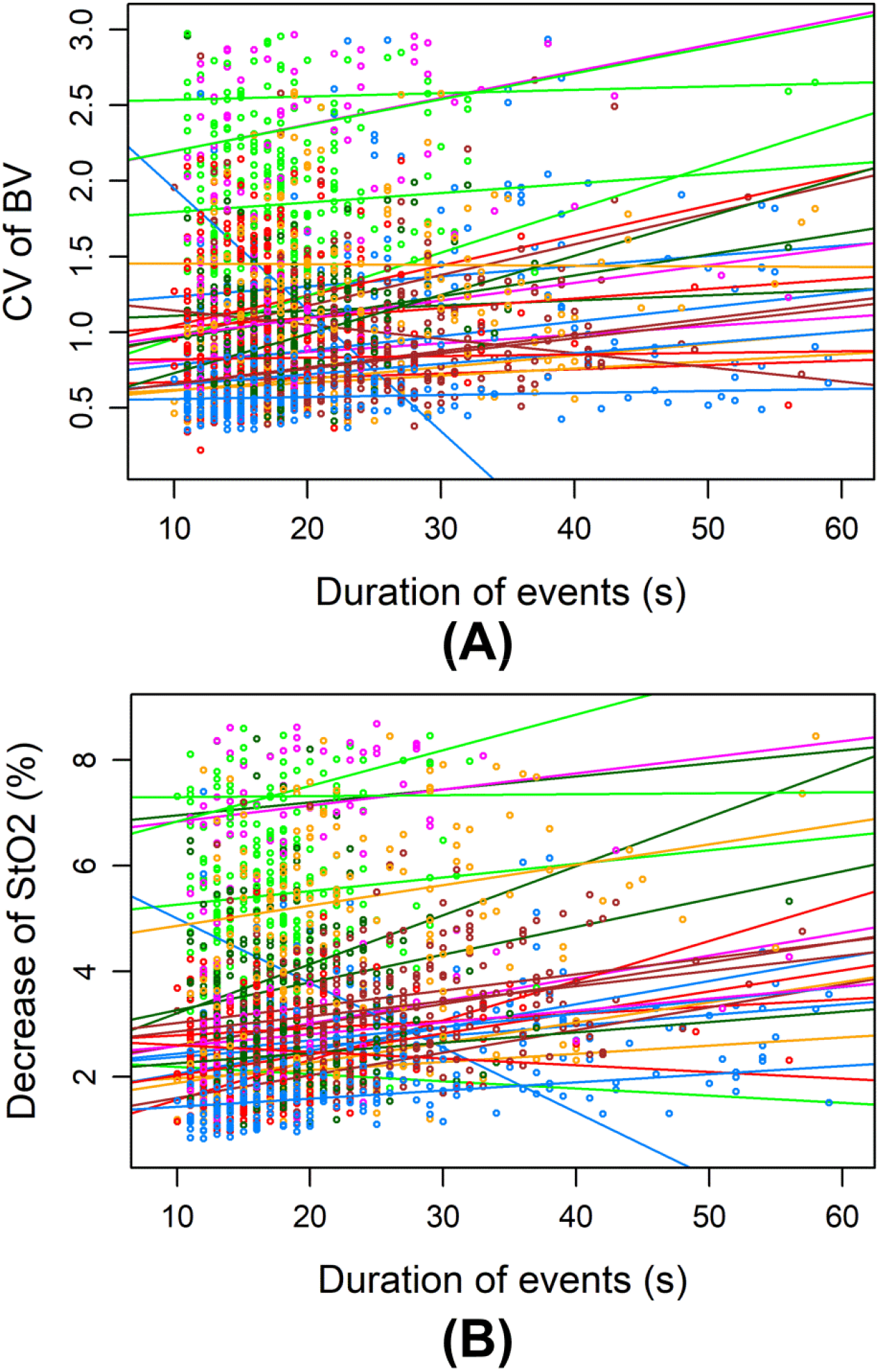
The changes in the coefficient of variation (CV) of cerebral blood volume (BV) and the decrease of cerebral tissue oxygen saturation (StO2) versus the duration of events in each patients. The data points of different colors indicate data from different patients, and the lines of different colors are the linear fitting of the data points. We only show the events of durations shorter than 60 seconds, considering that only a minority of events is longer than 60 seconds and they may be outliers that can bias the fitting trends.

## Discussion

Our study for the first time quantifies the relative effect sizes of different pathophysiological mechanisms contributing to the cerebral hemodynamic changes in local microvascular bed induced by obstructive sleep apneas and hypopneas. The strengths of our study are: 1) We use a stepwise increment CPAP titration protocol in which the first hour sleep without CPAP is served as a self-control in newly diagnosed untreated patients. The stepwise CPAP titration gradually opens the upper airway, increases ITP, reduces hypoxia and hypercapnia, reduces the numbers of apneas/hypopneas and arousals, and finally consolidates sleep. This protocol allows us to gradually control and identify the most relevant factors from many potential physiological factors in a systematic way, and our results are not affected by pre-treatment effect as our patients are newly diagnosed; 2) We simultaneously quantify the changes in both cerebral perfusion (i.e., changes in BV) and oxygenation (i.e., changes in StO2), thus providing better insights into the relationship between local cerebral metabolism and blood supply during apneas/hypopneas. Remarkably, we find that duration and the type of the respiratory events rather than AHI are the most significant covariates determining the cerebral hemodynamic changes in severe OSAS. Longer events induce stronger changes in cerebral perfusion and larger cerebral desaturation. Obstructive apneas lead to larger changes in cerebral BV and desaturation than hypopneas. In addition, brain activity (sleep stages, arousal and baseline cerebral oxygenation) as well as systemic parameter (i.e., HR and CPAP pressures) affect cerebral hemodynamics. Finally, our data demonstrate that CPAP is able to attenuate the cerebral hemodynamic changes triggered by the apneas/hypopneas.

Given that the event’s inherent features such as duration and the type of event, are the most significant variables after controlling for other covariates (these results are also valid in naive apneas/hypopneas at baseline, see supplemental Results), our findings have important scientific and clinical implications. Our results suggest that the duration and type of the event rather than AHI is a useful parameter indicating the severity of the cerebral damage in severe OSAS. Epidemiological studies investigating the cerebro-cardiovascular risks of OSAS should consider the length and type of respiratory events as predictors in the future. In clinical practice a summary of the length of respiratory events may be needed in traditional PSG report in addition to AHI. Patients with extreme long apneas/hypopneas may need to pay special attention and CPAP treatment should be highly recommended even if their AHI values are not high because a progressive increase in the events’ length may cause strong challenge of cerebrovascular autoregulation and an increased cerebrovascular risk. Our models also provide quantitative tools to assess the severity of cerebral hemodynamic changes. According to our model a longer duration of 5 seconds could induce 1.33×10^−2^ stronger (i.e., 2.66×10^−3^×5) changes in CV-BV but the obstructive apnea in general causes 19.53×10^−2^ more changes than hypopnea. Thus, a 15-second hypopnea will cause larger changes in cerebral perfusion than a 10-second hypopnea but smaller changes in cerebral perfusion compared to a 10-second obstructive apnea.

The contributions of different covariates to the cerebral hemodynamic regulations quantified by our models are reasonable and interpretable, and are consistent with each other. Higher HR during the apnea/hypopnea events may indicate relative higher sympathetic activation and less reduction in systemic blood supply [19, 38]. More systemic blood supply is available for the brain in these events compared to in the ones with lower HR, thus they trigger smaller changes in cerebral perfusion and StO2. This could consistently explain why we found higher baseline StO2 before the onsets of events could prevent the subsequent changes in cerebral perfusion and desaturation. Previous studies found that longer apnea events are associated with larger decrease in HR (i.e., smaller HR during the events) [38]. So it is reasonable that in our model the duration of the event and the mean HR within the event have contrary influences to the cerebral hemodynamics (see their fitted coefficients in Table 1 and 2).

The triggered cerebral hemodynamic changes are smaller in REM sleep compared to in NREM sleep, contrasting the existing data that during nocturnal REM sleep the obstructive apneas/hypopneas are associated with more severe peripheral oxygen desaturations [39-41]. One previous NIRS study also reported that the drops of cerebral StO2 induced by obstructive sleep apneas/hypopneas are larger in REM sleep compared to in NREM sleep [27]. The different results could be explained by: 1). In our stepwise incremental CPAP titration protocol the pressures are higher in REM sleep because REM sleep occurs mainly in the later period of sleep toward morning. So it is more likely that CPAP better restore the obstructive apneas/hypopneas in REM sleep compared to in NREM sleep. 2). The previous NIRS study [27] actually measured 13 patients during daytime nap without CPAP and only 4 patients had REM sleep. Most of their patients (8/13) had been on regular CPAP therapy before the study. So their results are unlikely to be comparable with ours, considering the different study protocols (i.e., nocturnal CPAP titration vs. normal daytime nap), different study populations (i.e., CPAP treatment naïve vs. regular CPAP treatment) and their small number of patients having REM sleep. 3). Cerebral oxygen saturation may be different from the peripheral one considering the unique cerebral CA mechanism. Previous studies found that the cerebral StO2 measured by NIRS keeps increasing over the whole night sleep [42], indicating the brain may have higher oxygen supply in REM sleep. Therefore, we may hypothesize that probably even without CPAP the naïve apneas/hypopneas may cause smaller desaturation in the brain in REM sleep compared to in NREM sleep after controlling the other covariates (i.e., length and type of event, HR, sleep position, etc.), in spite of the stronger desaturation in peripheral tissues. This hypothesis needs to be tested in the future studies.

Sleep position can influence the changes in cerebral perfusion but not cerebral oxygenation. This result can be explained by the gravity that influences the local cerebral blood supply and CA [43]. As our NIRS sensor is placed on the left forehead higher perfusion in the left hemisphere is measured when the patients sleep on their left side compared to on their right side or on their back. This result indicates that cerebral blood supply can be regulated by factors such as sleep position independent from cerebral oxygen consumption during apneas/hypopneas.

### Limitations and perspectives

There are several limitations in this study that need to be further improved in the future studies. First, the AHI is not a significant predictor, suggesting that the frequency of the apnea/hypopnea attacks is not relevant anymore for the brain damage in severe OSAS. This is probably because the CA functions of our patients have been already impaired, considering their age. Whether this is a common phenomenon also existing in mild and moderate OSAS patients is an interesting question needing further experimental studies. Second, the function of CA decreases with aging but age is not a significant predictor for changes in cerebral hemodynamics in our models. One possible explanation is that we only include patients with severe OSAS who need CPAP therapy, thus we could not include many young patients to cover a larger range of age. Third, we do not measure CO2 which may be a strong factor influencing cerebral hemodynamics because of the CPAP mask. Finally, we did not include central sleep apneas into analysis because it is difficult to distinguish the naïve ones from the treatment-emergent ones [44]. Future studies thoroughly studying the patterns of cerebral hemodynamics may be helpful to classify different central apneas and deepen our knowledge of the pathophysiological features/regulations of different types of sleep apneas and their hemodynamic consequences to the brain.

## Data Availability

The data are available from the authors on a reasonable request.

## Author contribution

Study design: ZZ, RK; Data acquisition: MQ; Data analysis: ZZ, GH, RK; Data interpretation: ZZ, RK; Paper writing: ZZ, RK.

## References

1. Senaratna CV, Perret JL, Lodge CJ, Lowe AJ, Campbell BE, Matheson MC, et al. Prevalence of obstructive sleep apnea in the general population: A systematic review. Sleep Med Rev. 2017;34:70–81. Epub 2016/08/29. doi: S1087-0792(16)30064-8 [pii] 10.1016/j.smrv.2016.07.002s. PubMed PMID: 27568340.

2. Heinzer R, Vat S, Marques-Vidal P, Marti-Soler H, Andries D, Tobback N, et al. Prevalence of sleep-disordered breathing in the general population: the HypnoLaus study. Lancet Respir Med. 2015;3(4):310–8. Epub 2015/02/16. doi: 10.1016/S2213-2600(15)00043-0 S2213-2600(15)00043-0 [pii]. PubMed PMID: 25682233; PubMed Central PMCID: PMC4404207.

3. Berry RB, Brooks R, Gamaldo CE, Harding SM, Lloyd RM, Marcus CL, et al. The AASM Manual for the Scoring of Sleep and Associated Events: Rules, Terminology and Technical Specifications. Darien, IL: American Academy of Sleep Medicine; 2017.

4. Redline S, Yenokyan G, Gottlieb DJ, Shahar E, O’Connor GT, Resnick HE, et al. Obstructive sleep apnea-hypopnea and incident stroke: the sleep heart health study. Am J Respir Crit Care Med. 2010;182(2):269–77. Epub 2010/03/27. doi: 10.1164/rccm.200911-1746OC. PubMed PMID: 20339144; PubMed Central PMCID: PMCPMC2913239.

5. Arzt M, Young T, Finn L, Skatrud JB, Bradley TD. Association of sleep-disordered breathing and the occurrence of stroke. Am J Respir Crit Care Med. 2005;172(11):1447–51. Epub 2005/09/06. doi: 10.1164/rccm.200505-702OC. PubMed PMID: 16141444; PubMed Central PMCID: PMCPMC2718439.

6. Yaggi HK, Concato J, Kernan WN, Lichtman JH, Brass LM, Mohsenin V. Obstructive sleep apnea as a risk factor for stroke and death. New Engl J Med. 2005;353(19):2034–41. doi: Doi 10.1056/Nejmoa043104. PubMed PMID: ISI:000233119600008.

7. Bassetti CL, Milanova M, Gugger M. Sleep-disordered breathing and acute ischemic stroke: diagnosis, risk factors, treatment, evolution, and long-term clinical outcome. Stroke. 2006;37(4):967–72. Epub 2006/03/18. doi: 10.1161/01.STR.0000208215.49243.c3. PubMed PMID: 16543515.

8. Furtner M, Staudacher M, Frauscher B, Brandauer E, Esnaola y Rojas MM, Gschliesser V, et al. Cerebral vasoreactivity decreases overnight in severe obstructive sleep apnea syndrome: a study of cerebral hemodynamics. Sleep Med. 2009;10(8):875–81. Epub 2009/02/10. doi: 10.1016/j.sleep.2008.09.011 S1389-9457(08)00311-0 [pii]. PubMed PMID: 19200779.

9. Hajak G, Klingelhofer J, Schulz-Varszegi M, Sander D, Ruther E. Sleep apnea syndrome and cerebral hemodynamics. Chest. 1996;110(3):670–9. Epub 1996/09/01. doi: S0012-3692(16)41069-X [pii]. PubMed PMID: 8797410.

10. Pizza F, Biallas M, Wolf M, Werth E, Bassetti CL. Nocturnal cerebral hemodynamics in snorers and in patients with obstructive sleep apnea: a near-infrared spectroscopy study. Sleep. 2010;33(2):205–10. Epub 2010/02/24. PubMed PMID: 20175404; PubMed Central PMCID: PMC2817907.

11. Zhang Z, Schneider M, Laures M, Qi M, Khatami R. The Comparisons of Cerebral Hemodynamics Induced by Obstructive Sleep Apnea with Arousal and Periodic Limb Movement with Arousal: A Pilot NIRS Study. Front Neurosci. 2016;10:403. Epub 2016/09/16. doi: 10.3389/fnins.2016.00403. PubMed PMID: 27630539; PubMed Central PMCID: PMCPMC5005379.

12. Schwarz EI, Furian M, Schlatzer C, Stradling JR, Kohler M, Bloch KE. Nocturnal cerebral hypoxia in obstructive sleep apnoea: a randomised controlled trial. Eur Respir J. 2018;51(5). Epub 2018/04/28. doi: 1800032 [pii]10.1183/13993003.00032-2018 13993003.00032-2018 [pii]. PubMed PMID: 29700104.

13. Ulrich S, Nussbaumer-Ochsner Y, Vasic I, Hasler E, Latshang TD, Kohler M, et al. Cerebral oxygenation in patients with OSA: effects of hypoxia at altitude and impact of acetazolamide. Chest. 2014;146(2):299–308. Epub 2014/05/09. doi: S0012-3692(15)48819-1 [pii] 10.1378/chest.13-2967. PubMed PMID: 24811331.

14. Balfors EM, Franklin KA. Impairment of cerebral perfusion during obstructive sleep apneas. Am J Respir Crit Care Med. 1994;150(6 Pt 1):1587–91. Epub 1994/12/01. doi: 10.1164/ajrccm.150.6.7952619. PubMed PMID: 7952619.

15. Tzeng YC, Willie CK, Atkinson G, Lucas SJ, Wong A, Ainslie PN. Cerebrovascular regulation during transient hypotension and hypertension in humans. Hypertension. 2010;56(2):268–73. Epub 2010/06/16. doi: HYPERTENSIONAHA.110.152066 [pii] 10.1161/HYPERTENSIONAHA.110.152066. PubMed PMID: 20547971.

16. Pizza F, Biallas M, Kallweit U, Wolf M, Bassetti CL. Cerebral Hemodynamic Changes in Stroke During Sleep-Disordered Breathing. Stroke. 2012;43(7):1951–3. doi: Doi 10.1161/Strokeaha.112.656298. PubMed PMID: ISI:000305882000046.

17. Durgan DJ, Bryan RM, Jr. Cerebrovascular consequences of obstructive sleep apnea. J Am Heart Assoc. 2012;1(4):e000091. Epub 2012/11/07. doi: 10.1161/JAHA.111.000091 jah366 [pii]. PubMed PMID: 23130152; PubMed Central PMCID: PMC3487354.

18. Somers VK, Dyken ME, Clary MP, Abboud FM. Sympathetic neural mechanisms in obstructive sleep apnea. J Clin Invest. 1995;96(4):1897–904. Epub 1995/10/01. doi: 10.1172/JCI118235. PubMed PMID: 7560081; PubMed Central PMCID: PMC185826.

19. Kohler M, Stradling JR. Mechanisms of vascular damage in obstructive sleep apnea. Nat Rev Cardiol. 2010;7(12):677–85. Epub 2010/11/17. doi: 10.1038/nrcardio.2010.145. PubMed PMID: 21079639.

20. Andreas S, Hajak G, von Breska B, Ruther E, Kreuzer H. Changes in heart rate during obstructive sleep apnoea. Eur Respir J. 1992;5(7):853–7. Epub 1992/07/01. PubMed PMID: 1499710.

21. Cipolla M, editor. Control of Cerebral Blood Flow: San Rafael (CA): Morgan & Claypool Life Sciences; 2009.

22. Zhang Z, Khatami R. A Biphasic Change of Regional Blood Volume in the Frontal Cortex During Non-rapid Eye Movement Sleep: A Near-Infrared Spectroscopy Study. Sleep. 2015. Epub 2015/03/13. doi: sp-00437-14 [pii]. PubMed PMID: 25761983.

23. Zhang Z, Khatami R. Predominant endothelial vasomotor activity during human sleep: a near-infrared spectroscopy study. Eur J Neurosci. 2014. Epub 2014/08/27. doi: 10.1111/ejn.12702. PubMed PMID: 25156240.

24. Fischer AQ, Taormina MA, Akhtar B, Chaudhary BA. The effect of sleep on intracranial hemodynamics: a transcranial Doppler study. J Child Neurol. 1991;6(2):155–8. Epub 1991/04/01. PubMed PMID: 1904461.

25. Nasi T, Virtanen J, Toppila J, Salmi T, Ilmoniemi RJ. Cyclic alternating pattern is associated with cerebral hemodynamic variation: a near-infrared spectroscopy study of sleep in healthy humans. PLoS One. 2012;7(10):e46899. Epub 2012/10/17. doi: 10.1371/journal.pone.0046899 PONE-D-12-14751 [pii]. PubMed PMID: 23071658; PubMed Central PMCID: PMC3468598.

26. Birzis L, Tachibana S. Local Cerebral Impedance and Blood Flow during Sleep and Arousal. Exp Neurol. 1964;9:269–85. Epub 1964/04/01. doi: 10.1016/0014-4886(64)90024-x. PubMed PMID: 14142795.

27. Valipour A, McGown AD, Makker H, O’Sullivan C, Spiro SG. Some factors affecting cerebral tissue saturation during obstructive sleep apnoea. Eur Respir J. 2002;20(2):444–50. Epub 2002/09/06. doi: 10.1183/09031936.02.00265702. PubMed PMID: 12212980.

28. Olopade CO, Mensah E, Gupta R, Huo D, Picchietti DL, Gratton E, et al. Noninvasive determination of brain tissue oxygenation during sleep in obstructive sleep apnea: a near-infrared spectroscopic approach. Sleep. 2007;30(12):1747–55. Epub 2008/02/06. PubMed PMID: 18246984; PubMed Central PMCID: PMC2276122.

29. Kainerstorfer JM, Sassaroli A, Tgavalekos KT, Fantini S. Cerebral autoregulation in the microvasculature measured with near-infrared spectroscopy. J Cereb Blood Flow Metab. 2015;35(6):959–66. Epub 2015/02/12. doi: 10.1038/jcbfm.2015.5 jcbfm20155 [pii]. PubMed PMID: 25669906; PubMed Central PMCID: PMC4640259.

30. Netzer NC. Impaired nocturnal cerebral hemodynamics during long obstructive apneas: the key to understanding stroke in OSAS patients? Sleep. 2010;33(2):146–7. Epub 2010/02/24. PubMed PMID: 20175397; PubMed Central PMCID: PMC2817901.

31. Liu H, Chance B, Hielscher AH, Jacques SL, Tittel FK. Influence of blood vessels on the measurement of hemoglobin oxygenation as determined by time-resolved reflectance spectroscopy. Med Phys. 1995;22(8):1209–17. Epub 1995/08/01. PubMed PMID: 7476706.

32. Iannetta D, Inglis EC, Soares RN, McLay KM, Pogliaghi S, Murias JM. Reliability of microvascular responsiveness measures derived from near-infrared spectroscopy across a variety of ischemic periods in young and older individuals. Microvasc Res. 2019;122:117–24. Epub 2018/10/08. doi: S0026-2862(18)30204-8 [pii] 10.1016/j.mvr.2018.10.001. PubMed PMID: 30292692.

33. Fantini S, Franceschini M-A, Maier JS, Walker SA, Barbieri BB, Gratton E. Frequency-domain multichannel optical detector for noninvasive tissue spectroscopy and oximetry. OPTICE. 1995;34(1):32–42.

34. Fantini S, Sassaroli A. Frequency-Domain Techniques for Cerebral and Functional Near-Infrared Spectroscopy. Front Neurosci. 2020;14:300. Epub 2020/04/23. doi: 10.3389/fnins.2020.00300. PubMed PMID: 32317921; PubMed Central PMCID: PMC7154496.

35. McLay KM, Nederveen JP, Pogliaghi S, Paterson DH, Murias JM. Repeatability of vascular responsiveness measures derived from near-infrared spectroscopy. Physiol Rep. 2016;4(9). Epub 2016/05/06. doi: 10.14814/phy2.12772 e12772 [pii] 4/9/e12772 [pii]. PubMed PMID: 27147496; PubMed Central PMCID: PMC4873629.

36. Nakagawa S, Schielzeth H. A general and simple method for obtaining R2 from generalized linear mixed-effects models. Methods in Ecology and Evolution. 2013;4(2):133–42. doi: 10.1111/j.2041-210x.2012.00261.x.

37. Xu R. Measuring explained variation in linear mixed effects models. Stat Med. 2003;22(22):3527–41. Epub 2003/11/06. doi: 10.1002/sim.1572. PubMed PMID: 14601017.

38. Zwillich CW. Sleep apnoea and autonomic function. Thorax. 1998;53 Suppl 3:S20–4. Epub 1999/04/08. PubMed PMID: 10193356; PubMed Central PMCID: PMCPMC1765913.

39. Findley LJ, Wilhoit SC, Suratt PM. Apnea duration and hypoxemia during REM sleep in patients with obstructive sleep apnea. Chest. 1985;87(4):432–6. Epub 1985/04/01. doi: 10.1378/chest.87.4.432. PubMed PMID: 3979129.

40. Haba-Rubio J, Janssens JP, Rochat T, Sforza E. Rapid eye movement-related disordered breathing: clinical and polysomnographic features. Chest. 2005;128(5):3350–7. Epub 2005/11/24. doi: 10.1378/chest.128.5.3350. PubMed PMID: 16304283.

41. Mokhlesi B, Finn LA, Hagen EW, Young T, Hla KM, Van Cauter E, et al. Obstructive sleep apnea during REM sleep and hypertension. results of the Wisconsin Sleep Cohort. Am J Respir Crit Care Med. 2014;190(10):1158–67. Epub 2014/10/09. doi: 10.1164/rccm.201406-1136OC. PubMed PMID: 25295854; PubMed Central PMCID: PMCPMC4299639.

42. Metz AJ, Pugin F, Huber R, Achermann P, Wolf M. Brain tissue oxygen saturation increases during the night in adolescents. Adv Exp Med Biol. 2013;789:113–9. Epub 2013/07/16. doi: 10.1007/978-1-4614-7411-1_16. PubMed PMID: 23852484.

43. Petersen LG, Ogoh S. Gravity, intracranial pressure, and cerebral autoregulation. Physiol Rep. 2019;7(6):e14039. Epub 2019/03/27. doi: 10.14814/phy2.14039. PubMed PMID: 30912269; PubMed Central PMCID: PMCPMC6434070.

44. Edwards BA, Malhotra A, Sands SA. Adapting our approach to treatment-emergent central sleep apnea. Sleep. 2013;36(8):1121–2. Epub 2013/08/02. doi: 10.5665/sleep.2862. PubMed PMID: 23904668; PubMed Central PMCID: PMCPMC3700705.

