## supplemental for "Is sleep apnea-hypopnea index relevant for impaired brain perfusion and desaturation in patients with severe obstructive sleep apnea syndromes?"

**Supplemental Material**

**Methods**

**Frequency-domain multi-distance (FDMD) near-infrared spectroscopy (NIRS)**

FDMD-NIRS (Imagent, ISS, Champaign IL, USA) measurements were conducted over the middle of left bicep muscle. Its light emitters, four laser diodes at 690-nm wavelength and four laser diodes at 830-nm wavelength were coupled into four light sources and were high frequency modulated at 110 MHz. The light can penetrate into the measured tissues with a depth of several centimeters when the four light sources were aligned and placed at 2 cm, 2.5 cm, 3 cm and 3.5 cm away from an optical fiber bundle connected to the photomultiplier tube detector.

Like other non-invasive optical methods measuring hemodynamic changes in human tissues such as photoplethysmography and other NIRS techniques (e.g., continuous wave NIRS), the basic principle of FDMD-NIRS is the modified Beer–Lambert law (MBLL) [1, 2]. In the original Beer–Lambert law the light extinction (i.e., the logarithm of the ratio of incident versus measured light) is proportional to the concentration of the absorber multiplied by the constant extinction coefficient for the particular absorber and the source-detector distance when light passes through a non-scattering but absorbing media [3]. However, biological tissues are highly scattering media and scattering will increase the path-length of light, increasing the probability of both light absorption and loss of light. Thus in MBLL the light attenuation due to scattering is taken into account by introducing the differential path-length factor (DPF). The real path-length of light in the media is then calculated as DPF multiplies the source-detector distance. DPF varies between different biological tissues and different individuals, and it also depends on other factors like light wavelength, age and gender [4, 5]. Its value increases with increasing scattering (i.e., the light can travel longer pathway in the tissues) and decreases with increasing absorption (i.e., the light is more likely to be absorbed soon after entering into the tissues) [6]. Thus DPF can be calculated using the following equation [4]:

$DPF=\frac{1}{2}\frac{\sqrt{3\mu_{s}^{'}}}{\sqrt{\mu_{a}}}$	(1)

where $\mu_{a}$ is the absorption coefficient and $\mu_{s}'$ is the reduced scattering coefficient of the measured biological tissue. Currently the majority of the commercially available NIRS devices are based on the simple continuous wave NIRS technique [5], in which the DPF uses a fixed value in the range of 3 to 6 because they cannot measure$\mu_{s}'$ . Thus these devices can only estimate the relative changes in the main absorbing chromophores in the measured biological tissues which are oxygenated hemoglobin (HbO2) and deoxygenated hemoglobin (HHb). Our Imagent system is currently the only commercially benchtop FDMD-NIRS device than can measure both the $\mu_{a}$and $\mu_{s}'$ in the measured tissues [6-8]. As illustrated in Fig.1, the light emitted from light source can be detected by detectors placed at different distances away from the source. The light intensity and modulation amplitude of the detected light decrease and phase delay occurs between the detected light and the source light due to the absorption and scattering. The detected light intensity and modulation amplitude are smaller but the phase delay is larger at the detector further away from the light source. The measured light intensity ($I\text{DC}$), modulation amplitude ($\text{I}\text{AC}$) and phase delay ( $phase$) from different light source-detector distances actually vary linearly [6, 7]. Therefore, to submit the measured$\text{I}\text{DC}$, $I\text{AC}$ and $phase$ to linear regression we can obtain the following equations derived from photon diffusion equation in a semi-infinite geometry [6, 8, 9]:

$ln\text{(}r^{2}\text{I}\text{AC}\text{)=}r\text{S}\text{AC}\text{ +}\text{C}\text{AC}$	(2)

$ln\text{(}r^{2}\text{I}\text{DC}\text{)=}r\text{S}\text{DC}\text{ +}\text{C}\text{DC}$	(3)

$phase=r\text{S}\text{phase}\text{ +}\text{C}\text{phase}$	(4)

Where $r$ is the known source-detector distance, $S\text{AC}$, $S\text{DC}$ and $S\text{phase }$are the slopes and $C\text{AC}$, $C\text{DC}$, $C\text{phase}$ are the intercepts. The linearity of the measured light signals is monitored by the R^2^ of the fitted linear regression. Combing any two of these three slopes (e.g., we chose$\text{S}\text{AC}$ and $S\text{phase}$ in the following equations) we can calculate $\mu_{a}$and $\mu_{s}'$ of the measured tissue [6, 8, 9]:

$\mu_{a}=-\frac{\omega}{2\nu}\frac{\text{S}\text{AC}}{S\text{phase}}\left( \frac{{S\text{phase}}^{2}}{{\text{S}\text{AC}}^{2}}+1 \right)^{-1/2}$	(5)

$\mu_{s}'=\frac{{\text{S}\text{AC}}^{2}}{3\mu_{a}}-\mu_{a}$	(6)

where $\omega/2\pi$ is the modulation frequency and $\nu$ is the velocity of light in the tissue. $\mu_{a}$ and $\mu_{s}'$ of both wavelengths can be calculated individually using the same equations (5) and (6). The DPF of the two wavelengths thus can be calculated using equation (1) and then the absolute values of HbO2 and HHb can be derived with the MBLL [1, 2, 6, 7]. The StO2 is then calculated as:

$StO2=100\times\frac{HbO2}{HbO2+HHb}$	(7)

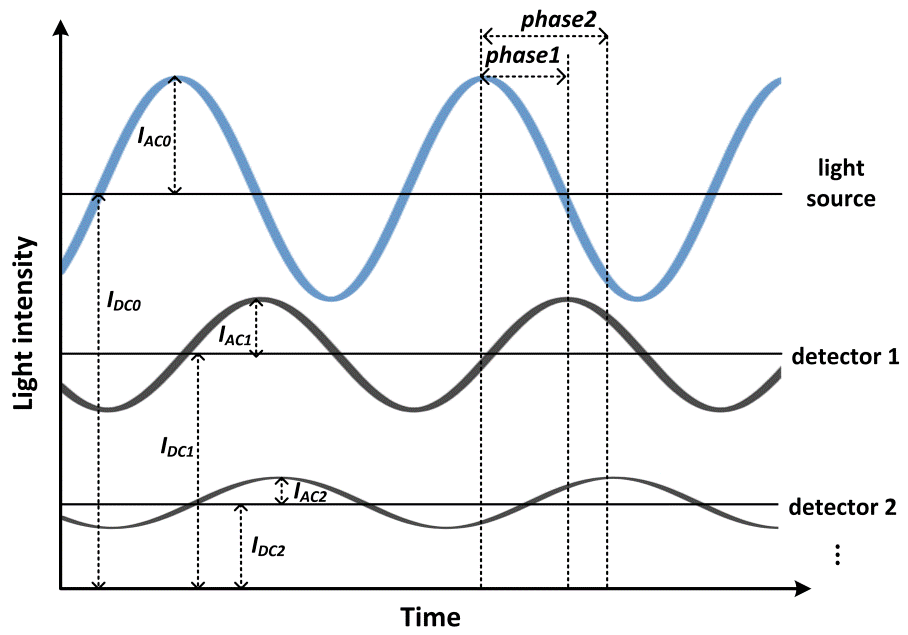

Figure 1. Frequency-domain multi-distance near-infrared spectroscopy measurement. The blue sine wave is the high frequency modulated light source. $I_{DC0}$, $I_{AC0}$ are its light intensity and modulation amplitude. The two black sine waves are the detected output light after travelling through the measured tissues. They are detected by detectors 1 and 2 placed at different distances away from the light source. The light intensities and modulation amplitudes of the two black sine waves are smaller than the ones of the light source and their phases are delayed, because of the absorption and scattering in the tissues. Detector 1 is closer to the light source than detector 2. Therefore, the light intensity $I_{DC1}$ and modulation amplitude $I_{AC1}$ detected at detector 1 are larger than the light intensity $I_{DC2}$ and modulation amplitude $I_{AC2}$ detected at detector 2. The phase delay $phase1$ at detector 1 is smaller than the phase delay $phase2$ at detector 2 because the light reaching detector 2 has travelled longer distance in the tissue. Similarly the light intensity and modulation amplitude will be further decreased and the phase delay will be further increased at the other detectors placed further away.

FDMD-NIRS system is currently well recognized as the most robust and reliable reference commercially available NIRS technology [7, 10, 11], considering its sophisticated mathematical frameworks calculating $\mu_{a}$ and$\mu_{s}'$ that can best estimate the real light propagation distance in the measured tissues based on derivations of the diffusion equation in complex geometries. The Imagent system used in this study has been approved by CE mark for research and its robustness, precision and accuracy of measuring HbO2/HHb/StO2 have been well validated in different physical blood-lipid models [6, 11, 12] and in vivo studies [13-15]. It has been used as a gold standard reference measurement of StO2 for the validations or calibrations of wearable [10] and portable [11, 16] NIRS oximeters such as the ones received FDA approval including INVOS 5100C (Medtronic, Dublin, Ireland)[17], FORE-SIGHT (CAS Medical Systems; Branford, CT, USA)[18] and SenSmart (Nonin Medical Inc., Plymouth, MN, USA)[19] .

**Pre-processing of FDMD-NIRS signals**

Before the start of every recording in the patients, the NIRS device was calibrated on optical phantom blocks. The reliability of FDMD-NIRS measurement depended on the linearity of the raw optical signals on distances, i.e., the linear dependence *R^2^* of modulated light amplitude and phase shift over the measure distances should be highly close to 1 in each light wavelength. The raw optical data were discarded if the *R^2^* was smaller than 0.95 in either modulation amplitude or phase shift in any wavelength. This step can exclude the poor quality data arising from improper probe-skin contact and the shunted light reaching the detectors without travelling through the tissue. Then the NIRS data were subjected to a low-pass (<0.08 Hz) zero-phase filter designed using Hanning window to remove the physiological noises including heart rate, respiratory noise and spontaneous slow hemodynamic oscillations[20, 21]. The filtered data were smoothed with moving average smooth method (robust locally weighted scatter plot smoothing [20, 22]). The StO2 values smaller than 30% or larger than 90% were discarded to exclude the potential movement artifacts or unreliable recordings, considering that the normal NIRS StO2 baseline value is between 50% and 80% [23, 24] and a drop up to 13% in StO2 is possible in some patients with brain dysfunction[25]. The whole event was excluded from further analysis if more than 20% of the data were discarded. In each patient if the mean BV of the event was more than three standard deviations away from the mean BV of all events, this event was defined as outlier and was excluded from further analysis.

**Results**

**The results of LMM and stepwise regression in native sleep apnea/hypopnea events at baseline without CPAP**

We used the same LMM and stepwise regression approach to further test if the result that the duration of events was the most significant predictor of changes in cerebral perfusion and desaturation in native sleep apnea/hypopnea events (i.e., at baseline without CPAP). The results of the final model for CV-BV and de-StO2 were summarized in e-Table 1 and e-Table 2 below, respectively.

1). The duration of the events and the types of events were the most significant predictors in both models after controlling the other covariates: longer events caused larger changes in cerebral perfusion and desaturation, and obstructive apneas triggered larger changes in cerebral hemodynamics than hypopneas.

2). The mean HR during the events and sleep positions were significant covariates influencing the changes in cerebral perfusion.

3). Baseline StO2 before the events was not a significant predictor for changes in cerebral perfusion at baseline but it was still significant for the cerebral desaturation (e-Table 2), which probably could be explained by the sleep initiation mechanisms that play an important role in cerebral perfusion in the first 1-hour baseline sleep. Our previous study showed increased cerebral perfusion after sleep onset using NIRS [26], suggesting that cerebral vasodilation may be a universal phenomenon at sleep onset process that is independent from (or less associated with) cerebral StO2.

4). Sleep stages did not influence changes in cerebral hemodynamics any more at baseline, which could be explained by the fact that in the first hour of sleep the patients rarely have REM sleep.

e-Table 1. The outcomes of the model of the CV-BV changes at baseline without CPAP.

|  | Estimate | Std. Error | t-value | P-value |
| --- | --- | --- | --- | --- |
| Duration of events | 0.01 | 0.0023 | 4.48 | <0.0001 |
| Mean HR within events | -0.0035 | 0.0015 | -2.27 | 0.024 |
| Apnea-Hypopnea | 0.33 | 0.046 | 7.213 | <0.0001 |
| Sleep positions | | | | |
| Right side | 0.14 | 0.096 | 1.48 | 0.13 |
| Supine | 0.289 | 0.083 | 3.46 | 0.0006 |

HR: heart rate. Sleep on left side is the reference for sleep on right side and on supine position. The conditional *R^2^* and *Ω^2^* of the model for CV-BV were 0.73 and 0.66, respectively.

e-Table 2. The outcomes of the model of the StO2 changes at baseline without CPAP.

|  | Estimate | Std. Error | t-value | P-value |
| --- | --- | --- | --- | --- |
| Duration of events | 0.043 | 0.0054 | 8.04 | <0.0001 |
| Baseline StO2 | -0.071 | 0.019 | -3.72 | 0.00022 |
| Apnea-Hypopnea | 0.42 | 0.11 | 3.98 | <0.0001 |

The conditional *R^2^* and *Ω^2^* of the model for CV-BV were 0.86 and 0.76, respectively.

**References**

1.	Villringer A, Chance B: **Non-invasive optical spectroscopy and imaging of human brain function**. *Trends Neurosci* 1997, **20**(10):435-442.

2.	Delpy DT, Cope M, van der Zee P, Arridge S, Wray S, Wyatt J: **Estimation of optical pathlength through tissue from direct time of flight measurement**. *Phys Med Biol* 1988, **33**(12):1433-1442.

3.	Beer: **Bestimmung der Absorption des rothen Lichts in farbigen Flüssigkeiten**. *Annalen der Physik* 1852, **162**(5):78-88.

4.	Scholkmann F, Wolf M: **General equation for the differential pathlength factor of the frontal human head depending on wavelength and age**. *J Biomed Opt* 2013, **18**(10):105004.

5.	Scholkmann F, Kleiser S, Metz AJ, Zimmermann R, Mata Pavia J, Wolf U, Wolf M: **A review on continuous wave functional near-infrared spectroscopy and imaging instrumentation and methodology**. *Neuroimage* 2014, **85 Pt 1**:6-27.

6.	Fantini S, Franceschini M-A, Maier JS, Walker SA, Barbieri BB, Gratton E: **Frequency-domain multichannel optical detector for noninvasive tissue spectroscopy and oximetry**. *Optical Engineering* 1995, **34**(1):32-42.

7.	Fantini S, Sassaroli A: **Frequency-Domain Techniques for Cerebral and Functional Near-Infrared Spectroscopy**. *Front Neurosci* 2020, **14**:300.

8.	Toronov V, Webb A, Choi JH, Wolf M, Safonova L, Wolf U, Gratton E: **Study of local cerebral hemodynamics by frequency-domain near-infrared spectroscopy and correlation with simultaneously acquired functional magnetic resonance imaging**. *Opt Express* 2001, **9**(8):417-427.

9.	Fantini S, Franceschini MA, Gratton E: **Semi-infinite-geometry boundary problem for light migration in highly scattering media: a frequency-domain study in the diffusion approximation**. *Journal of the Optical Society of America B* 1994, **11**(10):2128-2138.

10.	Farzam P, Starkweather Z, Franceschini MA: **Validation of a novel wearable, wireless technology to estimate oxygen levels and lactate threshold power in the exercising muscle**. *Physiological Reports* 2018, **6**(7):e13664.

11.	Kleiser S, Nasseri N, Andresen B, Greisen G, Wolf M: **Comparison of tissue oximeters on a liquid phantom with adjustable optical properties**. *Biomed Opt Express* 2016, **7**(8):2973-2992.

12.	Fantini S, Franceschini MA, Fishkin JB, Barbieri B, Gratton E: **Quantitative determination of the absorption spectra of chromophores in strongly scattering media: a light-emitting-diode based technique**. *Appl Opt* 1994, **33**(22):5204-5213.

13.	Stankovic MR, Maulik D, Rosenfeld W, Stubblefield PG, Kofinas AD, Drexler S, Nair R, Franceschini MA, Hueber D, Gratton E *et al*: **Real-time optical imaging of experimental brain ischemia and hemorrhage in neonatal piglets**. *J Perinat Med* 1999, **27**(4):279-286.

14.	Fantini S, Franceschini M, Gratton E, Hueber D, Rosenfeld W, Maulik D, Stubblefield P, Stankovic M: **Non-invasive optical mapping of the piglet brain in real time**. *Opt Express* 1999, **4**(8):308-314.

15.	Hallacoglu B, Sassaroli A, Wysocki M, Guerrero-Berroa E, Schnaider Beeri M, Haroutunian V, Shaul M, Rosenberg IH, Troen AM, Fantini S: **Absolute measurement of cerebral optical coefficients, hemoglobin concentration and oxygen saturation in old and young adults with near-infrared spectroscopy**. *J Biomed Opt* 2012, **17**(8):081406-081401.

16.	Kleiser S, Ostojic D, Andresen B, Nasseri N, Isler H, Scholkmann F, Karen T, Greisen G, Wolf M: **Comparison of tissue oximeters on a liquid phantom with adjustable optical properties: an extension**. *Biomed Opt Express* 2018, **9**(1):86-101.

17.	**U.S. Food and Drug Administration (FDA) 510(k) Database** [https://[www.accessdata.fda.gov/scripts/cdrh/cfdocs/cfpmn/pmn.cfm?ID=K971628](http://www.accessdata.fda.gov/scripts/cdrh/cfdocs/cfpmn/pmn.cfm?ID=K971628)]

18.	**U.S. Food and Drug Administration (FDA) 510(k) Database** [<http://www.accessdata.fda.gov/scripts/cdrh/cfdocs/cfpmn/pmn.cfm?ID=K094030>]

19.	**U.S. Food and Drug Administration (FDA) 510(k) Database** [https://[www.accessdata.fda.gov/scripts/cdrh/cfdocs/cfpmn/pmn.cfm?ID=K102715](http://www.accessdata.fda.gov/scripts/cdrh/cfdocs/cfpmn/pmn.cfm?ID=K102715)]

20.	Zhang Z, Schneider M, Laures M, Qi M, Khatami R: **The Comparisons of Cerebral Hemodynamics Induced by Obstructive Sleep Apnea with Arousal and Periodic Limb Movement with Arousal: A Pilot NIRS Study**. *Front Neurosci* 2016, **10**:403.

21.	Zhang Z, Khatami R: **Predominant endothelial vasomotor activity during human sleep: a near-infrared spectroscopy study**. *Eur J Neurosci* 2014, **40**(9):3396-3404.

22.	Cleveland WS, Devlin SJ: **Locally Weighted Regression - an Approach to Regression-Analysis by Local Fitting**. *J Am Stat Assoc* 1988, **83**(403):596-610.

23.	Quaresima V, Sacco S, Totaro R, Ferrari M: **Noninvasive measurement of cerebral hemoglobin oxygen saturation using two near infrared spectroscopy approaches**. *J Biomed Opt* 2000, **5**(2):201-205.

24.	Scheeren TW, Schober P, Schwarte LA: **Monitoring tissue oxygenation by near infrared spectroscopy (NIRS): background and current applications**. *J Clin Monit Comput* 2012, **26**(4):279-287.

25.	Al-Rawi PG, Kirkpatrick PJ: **Tissue oxygen index: thresholds for cerebral ischemia using near-infrared spectroscopy**. *Stroke* 2006, **37**(11):2720-2725.

26.	Zhang Z, Khatami R: **A Biphasic Change of Regional Blood Volume in the Frontal Cortex During Non-rapid Eye Movement Sleep: A Near-Infrared Spectroscopy Study**. *Sleep* 2015.
